# Determinants of Thyroid Dysfunction among Type 2 Diabetes Patients Attending Private Hospitals in Dire Dawa, Eastern Ethiopia

**DOI:** 10.1101/2022.02.03.22270379

**Authors:** Alemayehu Molla Tekalign, Fitsum Berhane Habte, Robel Mekonnen Yimer

**Author notes:** ***<u>Correspondening Author</u>*** Robel Mekonnen Yimer (MSc, Asst. Prof.), E-mail:- /.

## Abstract

**Purpose:** The purpose of this study is to capture the determinants of thyroid dysfunction in type 2 diabetes mellitus at private hospitals in Dire-Dawa, Eastern Ethiopia. Various studies found that thyroid dysfunction is common in type 2 diabetes and it is associated with poor blood glucose control and chronic diabetic complications responsible for morbidity and mortality. However, only limited studies have assessed determinants of thyroid dysfunctions among type 2 diabetes in Ethiopia.

**Methods:** Unmatched case-control study conducted in two private hospitals found in Dire Dawa town, between May - November 2021. A total of 92 type 2 diabetic patients with thyroid dysfunction as Cases and 183 type 2 diabetes patients with normal thyroid function as controls selected by purposive and systematic random sampling, respectively. Data collected by interview and using checklist, entered into SPSS version 22, and analyzed. Bivariate and multivariate logistic regression analysis employed to identify determinants of thyroid dysfunction with AOR and 95%CI. Independent t-test computed to observe significant mean difference of continuous variables. Variables with *P-value* < 0.05 declared as significant.

**Result:** Female gender predominate 65.9% of participants (p= 0.02). The mean glycated hemoglobin level among cases was 10.32 (± 2.4 SD) and 9.249 (± 2.3 SD) among controls, with significant difference (p=0.001). Similarly, the mean LDL cholesterol among cases 116.92 ±45.9 SD and control 102.34 ±43.97SD showed significant difference (p=0.016). Diabetic retinopathy and neuropathy, and ACEI/ARB also significantly associated with thyroid dysfunction (p< 0.05).

**Conclusion:** Female gender, lower educational level, and duration of diabetes associated with thyroid dysfunction. Most patients in both groups had unacceptably elevated HgA1c which need to be addressed. Thus, screening for thyroid dysfunction, especially the female gender, needs to be done.

## Introduction

Thyroid dysfunction (TD) is a condition characterized by an increase or decrease production of thyroid hormones with reciprocal change in pituitary secreted thyroid-stimulating hormone(TSH). All forms of TD - subclinical or clinical hypothyroidism or hyperthyroidism, mainly arise from pathology within the thyroid gland, although rarely they may originate from disorders of the hypothalamus or pituitary [1-2] and are associated with increased mortality [3].

The magnitude of TD is inconsistent in different parts of the world and relay on the level of iodine sufficiency, race, and population dynamics. The magnitude of overt hyperthyroidism ranges from 0.2% to 1.3%; hypothyroidism ranges from 1% to 2%, rising with age in iodine-sufficient parts of the world [4].

Studies have found a positive relationship between TD and type 2 diabetes mellitus (T2DM). There is also biochemical and clinical evidence that both endocrine diseases have shown to mutually influence each other [5-7] with reported 10-32% overall prevalence of TD in type 2 diabetes. Moreover, diabetes patients who have TD tend to have poorer glycemic control, an increased risk of lipid disorders, high blood pressure, and atherosclerosis which may accelerate diabetic vascular complications. A link between TD and microvascular complications and coronary artery diseases also reported [5, 8-9].

Despite an increasing global body of knowledge on the association of the two endocrine diseases, there is no clear guideline on screening T2DM patients for TD [10]. Moreover, predictors of TD in T2DM patients in different studies are inconsistent probably related to the difference in study design, characteristics of study subjects, and other conditions.

In Ethiopia, there is no nationwide prevalence or population-based data on the extent of TD, nor do we have any data on its prevalence in the diabetic population. Published or unpublished studies regarding TD among T2DM patients as well as predictors are also lacking. However, about 1.7 million adults aged 20–79 year old have had diabetes and the 2017 projected prevalence of diabetes in Ethiopia in these age groups was about 5.2% [11]. This indicates how big the problem is. Hence identifying determinants of TD in T2DM patients, contribution of TD for dysglycemia and chronic diabetic complications has public health importance.

When it comes to Dire Dawa, laboratory test for thyroid function and HgA1c is not currently done in public hospitals, and this leaves most T2DM patients not knowing their thyroid status and long term blood glucose control. In addition, there is knowledge gap among health professionals on the importance of TD in diabetes patients [12]. This study was aimed to identify predictors of TD in T2DM patients in Dire Dawa, Eastern Ethiopia. The findings have revealed important factors of TD and fill most of aforementioned gaps.

## Method and Materials

### Study area

The study was conducted in Dire Dawa city administration which is located at eastern part of Ethiopia, about 515km away from capital Addis Ababa. Dire Dawa comprises 453,000 population, of whom 227,406 (50.2%) are males and 225,594 (49.8) females.There are 6 hospitals (3 governmental, and 3 private), 15 Health center (8 urban and 7 rural), 34 health post, 56 primary, medium, and specialty clinic, and also 30 pharmacies, 42 drug vendor and 4 private hospital pharmacy found in the Administration.[13].

Bilal hospital is a private hospital owned by Ethio-American Physician, and started 15 years back. It provides inpatient and outpatient services in the disciplines of internal medicine, gynecology and obstetrics, and general surgical services. The hospital has well-established chronic disease follow-up clinics including diabetes, cardiovascular, and thyroid disorder. Patients served in the hospital include the surrounding regional states, neighboring Somalia, Somaliland state, and Djibouti. It has about 50 beds, 30 of which are dedicated to medical patients. Delt hospital is a tertiary care setup that incorporates all major medical specialties and gastroenterology subspecialty service. It provides inpatient for all major disciplines.

### Study Design and Period

Unmatched case-control study design employed and conducted between May 15 and July 15, 2021 in Bilal and DELT hospitals, Dire Dawa.

### Source and Study Population

The source populations consisted of T2DM patients who have follow up in selected private hospitals. Whereas, randomly selected T2DM patients with TD (cases) and without TD (controls) comprised the study population.

### Inclusion Criteria for Control

T2DM patients with at least 3 months follow up in selected hospitals, or later transferred to other hospitals in Dire Dawa who have normal TSH value (0.45-4.12 mU/L), or who was not taking treatment for thyroid dysfunction.

### Inclusion Criteria for Cases

T2DM patients who have TD based on abnormal TSH value (TSH < 0.45mU/L and TSH > 0.412 mU/L), or who was taking treatment for TD and with at least 3 months of follow up in selected hospitals, or later transferred to other hospitals in Dire-Dawa.

### Exclusion Criteria for Controls

T2DM patients who were taking or took amiodarone or underwent contrast imaging.

Patients with T2DM who are not tested for TSH value and their TD status remained unknown.

### Sample Size Determination

The sample size was determined using software Open-Epi, Version 3, open-source calculator--SSCC this web site https://www.openepi.com/SampleSize/SSCohort.htm, sample size calculations for unmatched case-control. By taking power of 80%, a cases to control ratio of 1:2, proportion of controls with exposure (0.435), proportion of cases with exposure (0.614) taking the proportion of elevated glycated hemoglobin level (> 7%) from Ogbanno et al 2019, the least Odds Ratio to be detected of 2.07 and 95% two-level confidence interval [14]. Accordingly, a total sample size of 275 (92 cases and 183 controls) taken.

### Sampling Procedure

A purposive sampling technique used to select cases as the cases are rare. Systematic random sampling method used to select controls at interval of every 2 control patients.

### Data Collection Procedure

Eligible study participants interviewed face to face using structured data collection tools. A data abstraction format used to collect information from participants’ medical records. The tool comprised information about socio-demographic, anthropometric characteristics of the patients, clinical variables taken from patient interviews, and a checklist to review patients’ medical records including laboratory, imaging variables, and drug regimens used by study participants. During an interview session, COVID-19 prevention measures strictly followed: well-ventilated room and maintaining 2 meter distance, data collectors and each interviewee wore face mask prior to interview, hand sanitizer and clean gloves used whenever needed.

### Study

#### Variables Dependent variable

- Thyroid dysfunction in type 2 diabetes patients

#### Independent variables

- Socio-demographic like age, gender, educational status, income, marital status, and occupation;
- Lifestyle factors including dietary preference, smoking, alcohol consumption and physical exercise;
- Knowledge and behavioral factors like adherence to diabetes self-management and knowledge of target blood glucose;
- Clinical and treatment factors such as clinical findings of chronic diabetic complications, clinical findings of thyroid dysfunction, echocardiography abnormality, drug regimens, laboratory tests including serum creatinine, blood lipid profile (total cholesterol, LDL, triglyceride, HDL, non HDL values), and hemoglobin level. Glycemic control was assessed by the level of glycated hemoglobin (HbA1c).

### Operational/ Standard Definition

#### Thyroid dysfunction

for this study, we use TSH as a marker of thyroid function. Based on literature TSH is the best and, often times, the only test needed. [15]

- Hypothyoridism is TSH > 4.12mU/L
- Hyperthyroidism if TSH values are < 0.45mU/L

#### Glycemic control

For this study, we categorized the study participants in to two groups based on the American Diabetes Association (ADA) recommendation [16]

- Good glycemic control: HbA1c< 7%
- Poor glycemic control: HbA1c > 7%

#### Glycosylated hemoglobin

Hemoglobin to which glucose is bound. The level of glycosylated hemoglobin is increased in the red blood cells of persons with poorly controlled diabetes mellitus. [67]

#### Diabetic nephropathy

is defined as a history of diabetes with the presence of albuminuria, impaired glomerular filtration rate, or both (eGFR <60ml/min/1.73m2) [17]

#### Estimated glomerular filtration rate (eGFR)

was calculated using the equation: 186× serum Cr–1.154 × age–0.203 × 0.742 (if female) ×1.233[17].

#### Diabetic neuropathy

was diagnosed based on abnormalities detected during neuropathic screening test (vibration sense testing, temperature sense, pain perception, monofilament testing, and examination of reflexes). [16]

#### Alcohol consumption

if reported consumption of alcohol twelve months before the study

#### Current tobacco smoke

is defined when a participant reported that he or she smoked tobacco products daily during the data collection period

#### Adherence to medication

if patients took all his/her anti-diabetic medication in last 7 days.

#### Adherence to blood glucose testing

if the patient measured his blood glucose for more than 3 days in last seven days.

#### Adherence to diet

If patients followed recommended diet for more than 3 days in last 7 days.

#### Adherence to exercise

If the patient follows recommended level of exercise for more than 3 days in last seven days.

### Data Processing and Analysis

The collected data coded and entered into Epi Data 4.2 software to minimize errors made while data entry. Then entered data transported to SPSS version 23.0 for analysis. Descriptive statistics like frequency, proportion, mean, and standard deviation employed to describe socio-demographic, clinical, and behavioral characteristics of patients. To identify predictors of thyroid dysfunction in diabetes, first, each independent variable with the outcome/dependent variable checked using bivariate logistic regressions. And those variables with p value <0.25 selected, imported, and adjusted using multivariate logistic regression with adjusted odds ratio (AOR), 95% CI. Variables with *P-value* < 0.05 declared statistically significant.

### Ethical Approval

Ethical clearance obtained from the Research Ethics Review Committee (RERC) of Dire-Dawa University. Permission granted by Dire Dawa Regional Health Bureau (DDRHB) and medical directors of Bilal and Delt hospitals. Informed consent signed and obtained from the participants after necessary explanation made about the purpose of the study, the risk and benefits, and also their rights of participating in the study. Interviews with subjects made with strict privacy and confidentiality was assured. The rights of respondents to refuse answering some or any questions respected. Those study participants who require treatment or modification of regimen have got feedback letters to the clinic where they do follow-up visits.

## Result

### Socio-demographic Comparison of cases and controls

A total of 275 subjects included in the final analysis: 92 were cases and the remaining 183 were controls. The mean age of subjects in cases was 53.6 ± 14.01 years and it was 55.65 ± 13.625 years in controls. The independent t-test showed no significant difference in the mean age between the case and control [t-value = -1.165; *p-value=* 0.245]. On the other hand, majority of participants were females accounting 72(78.2%) and 107(58.4%) among cases and controls, respectively. The difference in the proportion of gender between the two groups was tested and females are predominate which is statistically significant [AOR= 2.548 (1.15, 5.665); *p-value=* 0.02] (Table 1).

**Table 1:**
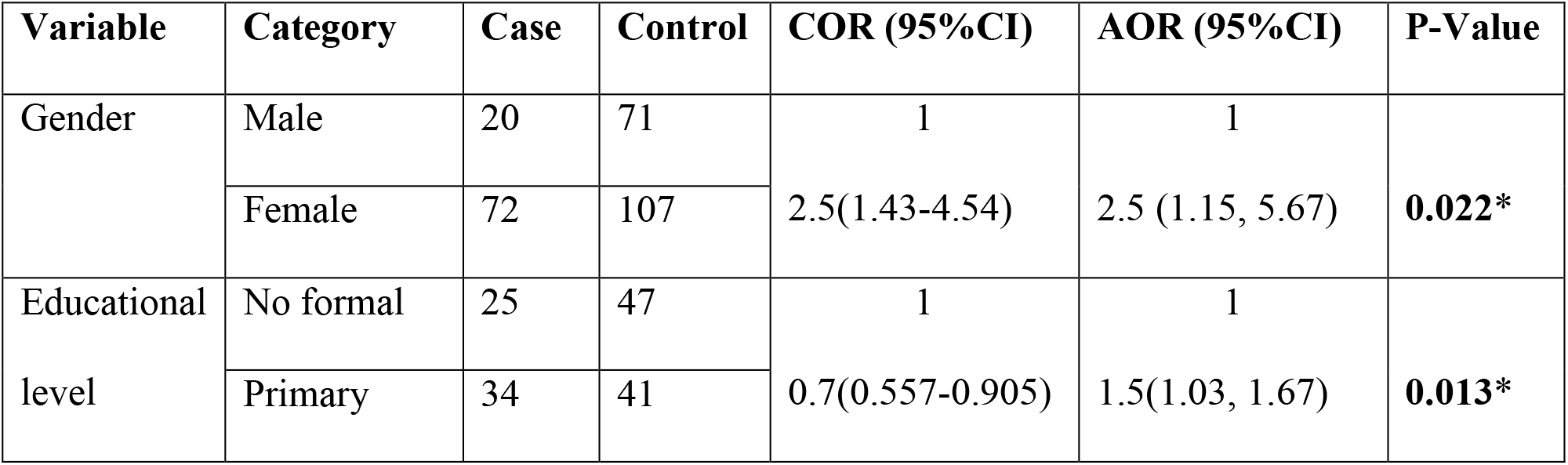
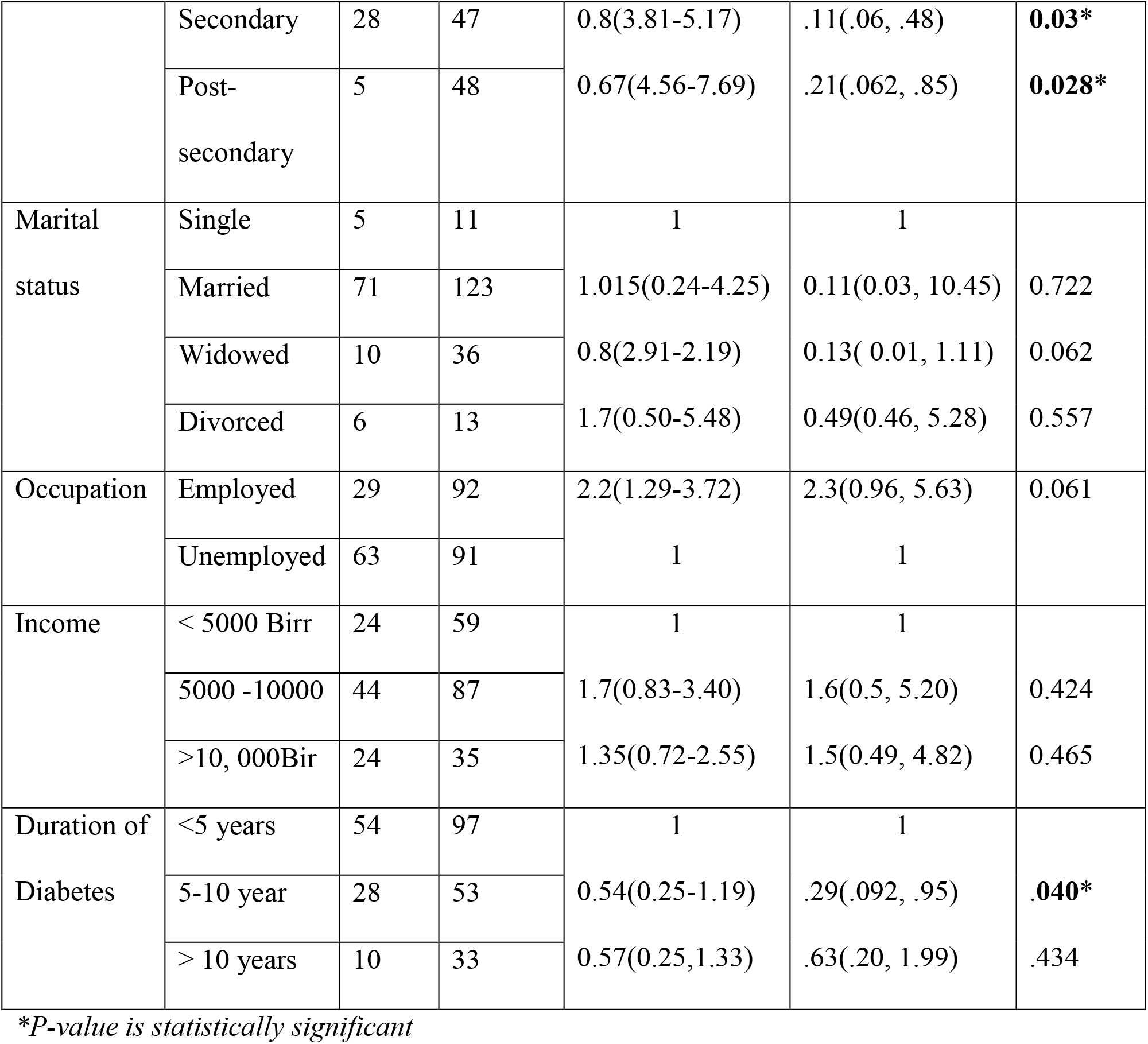
Comparison of Socio-demographic and Related Variables and Association with Dependent Variable.

Among the case, 25(27.2%) have no formal education, 34(37.0 %) have a primary education, and 28(30.4%) attended high school. Among the controls, 47(25.7%) have no formal education, 49.9(22.4%) and 49.9 (25.7%) attended primary and high school education. It was found that all forms of educational levels have significantly associated with thyroid dysfunction. Regarding employment status, 63(68.5%) of cases and almost half (49.7%) controls were unemployed. Even if it showed association in binary logistic regression [COR= 2.196; 95%CI (1.297-3.719)], failed when adjusted in multiple regression.

Out of the total 92 cases, 54(58,7%), 28(30.4%), and 10(10.9%) had a history of diabetes less than 5years, 5-10years, and more than 10years, respectively. Whereas, 101(55.2%), 50(27.3%), and 32(17.5%) controls had diabetes for less than 5years, 5-10years, and more than 10years, respectively. Having 5-10years of diabetes showed statistically significant association with TD [AOR = 0.95; 95%CI (0.9, .94); p = 0.04] (Table 1).

### Self-Care and Behaviors Related To Blood Glucose Control among Study Groups

It was found that 67% of cases consume a healthy diet more than 3 times per week including fruit, vegetables, high fiber diet, and low unsaturated fat; 72% of controls also do so. In addition, adherence to diabetic diet eating plan adequately practiced in 64% of cases and 72% of controls. However, both variables showed no significant association with TD (p>0.05) (Table 2).

**Table 2:** Self-Care and Behaviors Related To Blood Glucose Control among Study Groups.

| Variable | Category | Case | Control | COR (95%CI) | AOR (95%CI) | P-Value |
| --- | --- | --- | --- | --- | --- | --- |
| Exercise | <3x/wk | 66 | 123 | 1.2(0.71, 2.1) | 1.8(0.76, 4.30) | 0.182 |
|  | >3x/wk | 26 | 60 | 1 | 1 |  |
| SMBG | <3x/wk | 103 | 171 | 0.4(0.22, .73) | 0.55(0.21, 1.46) | 0.237 |
|  | >3x/wk | 30 | 26 |  |  |  |
| Smoking | Yes | 6 | 11 | 1.1(0.39, 3.05) | 0.91(0.31, 2.67) | 0.869 |
|  | No | 86 | 172 | 1 | 1 |  |
| Alcohol | Yes | 3 | 9 | 1.5(0.41, .80) | 1.0(0.99-1.01) | 0.741 |
|  | No | 89 | 174 | 1 | 1 |  |
| Hypertension | Yes | 49 | 90 | 1.2 (0.97, 1.94) | 0.9(0.44, 1.93) | 0.830 |
|  | No | 43 | 93 | 1 | 1 |  |
| Healthy diet | <3x/wk | 30 | 62 | 0.73( .42,1.3) | 1.2(.17, 8.74) | .855 |
|  | >3x/wk | 48 | 135 | 1 | 1 |  |
| Following | <3x/wk | 33 | 52 | 1.4(.82, 2.4) | .78(.11, 5.66) | .806 |
| dietary plan | >3x/wk | 59 | 131 | 1 | 1 |  |
| Unhealthy | <3x/wk | 49 | 121 | .58( .35, .97) | .66(.29, 1.49) | .318 |
| diet | >3x/wk | 43 | 62 | 1 | 1 |  |

*\*P-value is statistically significant*

Regarding an unhealthy diet such as high-fat food, full-fat dairy products, 46.7% of cases and 29.5 % of controls consume more than 3 times a week in the last 7 days. Although association between thyroid dysfunction and unhealthy diet observed on bivariate logistic regression, it failed when adjusted on multivariate analysis (p= 0.318). About 28.3% of cases and 32.8% controls involved at least 30 minutes of exercise 3 times per week.

Adherence to diabetes medication is also requested, 96% of cases and controls toke their medications at least 5 days per week. Only 6 of the 92 cases and 11 of the 183 controls have a smoking history. With the same token 3 of the cases and 9 of the controls have a history of alcohol intake, and 49(53%) of cases and 90(49.2%) of controls have hypertension diagnosed before and/or during this study. However, none of them showed significant association with TD.

### Glycemic Control and Thyroid Dysfunction

The mean glycated hemoglobin level of subjects in cases was 10.32 ± 2.4 % and it was 9.249 ± 2.3 in controls. The difference in the HbA1c between the two groups showed statistically significant as analyzed by an independent t-test [t= 3.518; df= 273; p= 0.001; 95% C.I (0.47, 1.67)]. The mean total cholesterol in cases (193.5 ± 49.47) was higher than the controls (184.35 ±53.7). In contrary, the mean of triglycerides and high-density lipoprotein (HDL) was higher for controls than the cases, yet no significant association observed. However, the mean of low-density lipoprotein (LDL)-cholesterol between cases (116.92 ±45.9) and controls (102.34 ±43.97) showed statistically significant difference in the means [(t= 2.420, df= 241, p= 0.016)] (Table 3).

**Table 3:** **Comparison between Participants with Thyroid Dysfunction and Euthyroid in Type 2 Diabetes Patients Regarding the Different Clinical Parameters**.

| Variable | Group | N | Mean | Std. Deviation | T | Significance | 95 % confidence interval |  |
| --- | --- | --- | --- | --- | --- | --- | --- | --- |
| Age | Case | 92 | 53.6 | 14.1 | -1.16 | 0.24 | -5.5 | 1.4 |
|  | Control | 183 | 55.65 | 13.6 |  |  |  |  |
| HbA1c level | Case | 92 | 10.3207 | 2.41531 | 3.518 | 0.001* | 0.47 | 1.167 |
|  | Control | 183 | 9.2497 | 2.36522 |  |  |  |  |
| Total Cholesterol | Case | 88 | 193.50 | 49.472 | 1.337 | 0.182 | 4.319 | 22.8 |
|  | Control | 175 | 184.35 | 53.697 |  |  |  |  |
| Triglyceride | Case | 88 | 160.83 | 93.772 | -.459 | 0.647 | -29.1 | 18.1 |
|  | Control | 173 | 166.33 | 90.393 |  |  |  |  |
| HDL | Case | 84 | 48.46 | 12.159 | 1.129 | 0.260 | -18.0 | 4.9 |
|  | Control | 159 | 55.04 | 52.643 |  |  |  |  |
| LDL | Case | 84 | 116.92 | 45.911 | 2.420 | 0.016* | 2.214 | 26.44 |
|  | Control | 159 | 102.34 | 43.972 |  |  |  |  |

\*P-value is statistically significant; HDL= high-density lipoprotein; LDL= low-density lipoprotein

### Thyroid Dysfunction and Diabetic Complications

On bivariate logistic regression analysis, diabetic retinopathy, diabetic foot, and diabetic neuropathy associated with thyroid dysfunction. After controlling the potential confounding factor including glycemic level, multivariate logistic regression identified diabetic neuropathy and retinopathy are independently associated with thyroid dysfunction. Accordingly, the odds of having thyroid dysfunction in patients with diabetic retinopathy is 9.3 times higher than diabetic patients with no retinopathy [(AOR= 9.334; 95%C.I. (2.049-42.51)]. Similarly, the odds of having an abnormal thyroid function test were 3.5 times higher in diabetic sensory neuropathy patients than patients without [(AOR= 3.523; 95 C. I. 1.640, 7.571)]. However, coronary artery disease and diabetic nephropathy have no association with thyroid dysfunction (Table 4).

**Table 4:** Comparison of Groups Concerning Diabetic Complication and Thyroid Dysfunction.

| Variable | Category | Case | Control | COR(95%CI) | AOR(95%CI) | P-Value |
| --- | --- | --- | --- | --- | --- | --- |
| Diabetic foot | Yes | 14 | 9 | 3.7(1.63, 8.4) | 3.5 (.79, 15.04) | 0.097 |
|  | No | 78 | 174 | 1 | 1 |  |
| Diabetic neuropathy | Yes | 46 | 41 | 1.92(1.07, 3.42) | 3.3(1.19, 8.92) | 0.021* |
|  | No | 46 | 142 | 1 | 1 |  |
| Diabetic retinopathy | Yes | 27 | 5 | 6.2(6.9, 10.5) | 9.3(2.05, 42.51) | 0.04* |
|  | No | 65 | 178 | 1 | 1 |  |
| Nephropathy | Yes | 19 | 34 | 0.03(0.02, 0.05) | 0.75(0.29, 1.99) | 0.571 |
|  | No | 72 | 148 | 1 | 1 |  |
| CAD | Yes | 31 | 70 | 1.2(0.72, 2.08) | 1.04(0.47, 2.28) | 0.931 |
|  | No | 61 | 113 | 1 | 1 |  |

\*P-value is statistically significant; CAD = Coronary Artery Disease

### Thyroid Dysfunction and Medications

The majority of patients, 67% of cases and 72.8% of controls took metformin. Whereas Sulfonuria is less used: (19%) of cases versus (24%) of controls. Insulin as a sole hypoglycemic agent used only in 9.7% of cases and 15.8% of controls; insulin with other oral hypoglycemic agents used in 20.6% of cases versus 14.8% of controls. However, none of the studied diabetic medications had statistically significant association (see Table 5).

**Table 5:** Comparison of Groups Related to Thyroid Dysfunction Associated with Drugs.

| Variable | Category | Case | Control | COR(95%CI) | AOR(95%CI) | P-Value |
| --- | --- | --- | --- | --- | --- | --- |
| METFORMIN | Yes | 67 | 123 | 1.3(.752- 2.27) | 1.3(0.56-2.69) | 0.605 |
|  | No | 25 | 60 | 1 | 1 |  |
| SULFONUREA | Yes | 18 | 44 | 0.77(.42-1.42) | 0.6(0.25- 1.52) | 0.289 |
|  | No | 74 | 139 | 1 | 1 |  |
| SGLT 2 | Yes | 3 | 6 | .99(.24-4.07) | 1.8 (.33-10.54) | 0.484 |
|  | No | 89 | 177 | 1 | 1 |  |
| INSULIN | Yes | 19 | 27 | 0.58 (.26-1.27) | 0.36(.11-1.16) | 0.90 |
|  | No | 73 | 155 | 1 | 1 |  |
| STATIN | Yes | 56 | 103 | 1.21(.73-2.01) | 0.8(.37-1.74) | 0.57 |
|  | No | 36 | 80 | 1 | 1 |  |
| ACE/ARB |  | 34 | 39 | 2.2(1.25-3.76) | 2.5(1.10- 5.73) | 0.028* |
|  |  | 58 | 144 | 1 | 1 |  |
| ANTIPLATE |  | 26 | 47 | 1.14(0.65-2.0) | 1.15(0.49-2.69) | 0.751 |
|  |  | 66 | 136 | 1 | 1 |  |

\*P-value is statistically significant; SGLT 2= Sodium-Glucose Co-Transporter 2
ACE/ARB= Angiotensin-Converting Enzyme Inhibitor/ Angiotensin II Receptor Blocker

Lipid lowering agent statins used in large proportions by both subjects: 60% cases and 56% controls. Yet its was not associated with the dependent variable. Angiotensin-converting enzyme inhibitor or angiotensin II blocker has been used in 34(36.9%) of cases and 39(21.3%) control patients. And this showed statistically significant association [AOR 2.16; 95% CI (1.247, 3.758); p= 0.005].

## Discussion

DM and thyroid disorders are the two common endocrinopathies seen in the adult population. Studies have shown a very high prevalence of thyroid dysfunction in type 2 DM. Insulin and thyroid hormones are intimately involved in cellular metabolism and thus excess or deficit of either of them may result in the functional derangement of the other [18-23]. This study assesses, the association of glycemic control with thyroid dysfunction, predictors of thyroid dysfunction, and association of thyroid dysfunction with chronic diabetic complications.

### Glycemic Control and Thyroid Dysfunction

In this study degree of blood glucose control in diabetes patients with thyroid dysfunction is poorer than controls as the mean glycated hemoglobin level was 10.32 ± 2.4% and it was 9.249 ± 2.3% in controls. The difference is statistically significant (p= 0.001) which underlines the association of chronic hyperglycemia and thyroid dysfunction. A similar case-control study done by Alsolami showed elevated HbA1c levels linked with thyroid dysfunction [24]. Contrary to our finding, Diez et al and Mahalingam, et al in their studies found no association [22-23, 25]. The association of hyperglycemia with thyroid dysfunction may be due to the adverse effects of chronic hyperglycemia on the hypothalamic-pituitary axis where it blunts or abolishes the nocturnal TSH peak [20, 14, 26]

As insulin resistance is thought to be central in the pathophysiology of the development of thyroid dysfunction, the duration of diabetes has been considered to be an important risk for thyroid dysfunction (more than 5 years) mainly in hypothyroidism. Our finding revealed that T2DM patients with 5 to 10 years duration of diabetes had higher odds of developing thyroid dysfunction than less duration. However, there is no association in this study [27, 14, 26]

### Socio-Demographic Predictors of Thyroid Dysfunction

In this study, females formed 65% of participants, and being female has 2.5 times the odds of having thyroid dysfunction than their male counterparts. This female preponderance is also demonstrated by Ogbonna (2019) [28], 60% of participants were females as well as studies done by Ghazali et al (2010) [21].

Both thyroid dysfunction and T2DM are among advanced age groups. In our study, the mean age of cases was 53.6(±14.01) years and 55.65(±13.625) years for controls. This was comparable with the report by Alolami where 55.7 years and 50.2 years the mean age of cases and controls, respectively. However, Ogbonna reported higher mean age of 57.5 years for cases and 57.7 years for controls. Despite advanced mean age of both groups, there was no significant difference observed which is in line with several studies [25, 29, 26].

This study also revealed that T2DM patients with just primary education had 1.5 times the odds of having thyroid dysfunction. On the other hand, T2DM patients who have secondary education and post-secondary education had less likely to have TD (p= 0.02) and (p= 0.028), this implies better educational levels being protective. This is logical since a better educational level may yield better blood glucose control which is associated with normal thyroid function. However, the Brazilian Longitudinal Study of Adult Health (ELSA-Brasil), showed highly educated patients have a higher frequency of thyroid disorder [29].

### Cardiovascular Risks and Thyroid Dysfunction in Type 2 Diabetes

In studies, thyroid dysfunction is considered a cardiovascular risk; one of the reasons is the presence of associated hyperlipidemia. Hypercholesterolemia, especially in hypothyroidism, probably results from reduced catabolism of lipoproteins, a phenomenon that may be explained by a decreased expression of lipoprotein receptors. In this study, level of LDL was associated with TD with mean LDL value of 116.92 (±45.9) for cases and 102.34 (±43.97) for control. This will predispose for the acceleration of atherosclerosis and early development of cardiovascular complications [28, 26]. Even if hypertension is common in diabetes and thyroid dysfunction, this study did not find an association with thyroid dysfunction. Hypertension as a predictor in several studies is inconsistent [20, 14].

### Diabetic Complications in Patients with Thyroid Dysfunction

Since TD is associated with poor blood glucose control, it is expected to worsen and accelerate the development of chronic complications. The effect of TD in aggravating microvascular complications of T2DM has been studied with inconsistent findings. In this study, T2DM patients with TD demonstrated strong association with diabetes retinopathy (p= 0.04). This was in line with the study done by Mohammed et al., subclinical hypothyroidism was associated with a higher frequency of microvascular (diabetic retinopathy) [26].

Moreover, significant association of diabetic neuropathy with thyroid dysfunction also observed (p= 0.021), but diabetic nephropathy did not associated. In contrary to our findings, studies done in Saudi Arabian, in Kuwait and Nigeria showed that T2DM patients with TD are more likely to suffer from nephropathy (P<0.001) [25, 14], but found no significant association with neuropathy and retinopathy [25, 14, 26].

It is known that hypothyroidism and T2DM are independently linked to the development of coronary artery disease. Studies reported thyroid dysfunction of all spectrums to have an association with coronary artery disease. However, this study did not, which could be due to the relatively small sample size. Further study, probably a prospective cohort study is needed to elucidate the presence and strength of association [30-31]. As a result, there is a general trend of increased complications in patients with thyroid dysfunction. This could imply that the coexistence of both thyroid dysfunction and T2DM could alter patients’ health conditions and metabolic control, and might also increase mortality. This could also decrease the quality of life for those patients if not detected early.

### Use of Medications and Thyroid Dysfunctions

Thyroid function is influenced by several drugs. Anti-diabetic drugs also incriminated in thyroid hormonal homeostasis. Metformin has a well-studied influence in lowering TSH, goitrogenic effect, Sulphonurias has reported higher incidence of hypothyroidism, and Insulin also increases the secretion of TRH and TSH [32]. This study has not found any association among anti-diabetic medications and statin with thyroid dysfunction. However, there is a significant association with angiotensin-converting enzyme inhibitors and angiotensin receptor blocking drugs (ACEI, ARB) (p=0.028). It can be related to the central effect of these drugs suppressing TSH secretion. Regardless, more study is necessary to justify its association, since no study has discovered by the investigator.

## Conclusion

Thyroid dysfunction in T2DM patients is a common condition associated with poor blood glucose control and chronic diabetic complications: diabetic retinopathy and diabetic neuropathy.

Diabetes patients with thyroid dysfunction have higher lipid levels (LDL cholesterol) than patients without thyroid dysfunction; as a result T2DM patients with TD are at higher risk of cardiovascular diseases. Female gender, lower educational level, and drugs are associated with thyroid dysfunction. Majority of patients, both in cases and controls groups, have unacceptably elevated glycated hemoglobin levels which need to be addressed to prevent morbidity and mortality. Thus, screening for thyroid dysfunction in T2DM patients, especially the female gender, needs to be done.

## Recommendation

### *Regional health bureau* should

- Consider to avail hormone test machine and reagents required for thyroid function test and glycated hemoglobin level at public health facilities.
- Create awareness among health professionals and to the public as diabetes is a public health problem in the region and significant portions of these populations are expected to have thyroid dysfunction.
- Attention should be given to enhance knowledge the population as this study showed increased level of education may prevent development of thyroid dysfunction

### To health facilities

- Health facilities should take the responsibility of screening diabetes patients for thyroid dysfunction specially women;
- Should give due attention to treat patient to the target following guideline
- Health facilities should provide adequate education

### To the patients

- Patients should know rationale of good glycemic control and be advised to adhere to medication and follow up and should be warned the consequence of persistent hyperglycemia and presence of thyroid dysfunction

### To researchers

Longitudinal study is essential to assess predictors over time; the reason for inconsistent association of chronic complication; and to further analyze effects of medication on thyroid function tests such as angiotensin converting enzymes inhibitors or angintenin blockers.

## Data Availability

All relevant data are within the manuscript and its Supporting Information files.

## Acknowledgment

We acknowledge Dire Dawa University, College of Medicine and Health Science, Department of Public Health facilitating to conduct this study. Our heartfelt gratitude goes to all study participants, all data collectors and health professionals in both hospitals for their unreserved help and facilitation. Finally, we would like to appreciate Dr. Kedir Teji, Sister Hanna Lambero and Nebiyu Damtew for their unreserved technical assistance.

## Notes

### Competing Interest Statement

The authors have declared no competing interest.

### Funding Statement

The authors received no specific funding for this work

### Author Declarations

Ethical approval was received from Dire Dawa University, Institutional Review Board.

### Summary of Updates

Author's last name updated

## References

1. Persani, L. Central hypothyroidism: pathogenic, diagnostic, and therapeutic challenges. J Clin Endocrinol Metab. 2012;97:3068–3078. doi:10.1210/jc.2012-1616

2. Hadlow NC, Rothacker KM, Wardrop R, Brown SJ, Lim EM, Walsh JP. The relationship between TSH and free T4 in a large population is complex and nonlinear and differs by age and sex. J Clin Endocrinol Metab. 2013 Jul;98(7):2936–43. doi: 10.1210/jc.2012-4223. Epub 2013 May 13. PMID: 23671314.

3. Laulund AS, Nybo M, Brix TH, Abrahamsen B, Jørgensen HL, Hegedüs L. Duration of thyroid dysfunction correlates with all-cause mortality. the OPENTHYRO Register Cohort. PLoS One. 2014;9(10):e110437. Published 2014 Oct 23. doi:10.1371/journal.pone.0110437

4. Taylor, P., Albrecht, D., Scholz, A. l. Global epidemiology of hyperthyroidism and hypothyroidism. Nat Rev Endocrinol 14, 301–316 (2018). https://doi.org/10.1038/nrendo.2018.18

5. Chaker, L., Bianco, A. C., Jonklaas, J. & Peeters, R. P. Hypothyroidism. Lancet, doi:10.1016/s0140-6736(17)30703-1 (2017).

6. Wang, “The relationship between type 2 diabetes mellitus and related thyroid diseases,” Journal of Diabetes Research, vol. 2013, Article ID 390534, 9 pages, 2013. [5]

7. Al-Geffari, N. A. Ahmad, A. H. Al-Sharqawi, A. M. Youssef, D. Alnaqeb, and K. Al-Rubeaan, “Risk factors for thyroid dysfunction among type 2 diabetic patients in a highly diabetes mellitus prevalent society,” International Journal of Endocrinology, vol. 2013, Article ID 417920, 6 pages, 2013.

8. Osei Sarfo-Kantanka, Eunice Oparebea Ansah, Ishmael Kyei, Nana Ama Barnes, Causes and predictors of mortality among Ghanaians hospitalised with endocrine disorders, International Health, Volume 12, Issue 2, March 2020, Pages 107–115, https://doi.org/10.1093/inthealth/ihz038

9. Ladenson, P. A. Singer, K. B. A, “American thyroid association guidelines for detection of thyroid dysfunction,” Archives of Internal Medicine, vol. 160, no. 11, pp. 1573–1575, 2000.

10. Brenta, S. Danzi, and I. Klein, “Potential therapeutic applications of thyroid hormone analogs,” Nature Clinical Practice Endocrinology and Metabolism, vol. 3, no. 9, pp. 632–640, 2007.

11. Centers for Disease Control and Prevention, National DiabetesStatistics Report, Atlanta, GA, USA, Centers for DiseaseControl and Prevention, US Dept of Health and HumanServices, 2017

12. Mengesha, M.M., Roba, H.S., Ayele, B.H. Level of physical activity among urban adults and the socio-demographic correlates: a population-based cross-sectional study using the global physical activity questionnaire. BMC Public Health 19, 1160 (2019). https://doi.org/10.1186/s12889-019-7465-y

13. Ethiopian Government Portal. The Dire Dawa administrative council. Ethiopian Government Portal. 2012 [cited 2016 Jan 9]; Available at: URL:http://www.ethiopia.gov.et/dire-dawa-city-administration.

14. Olmos RD, Figueiredo RC, Aquino EM, Lotufo PA, Bensenor IM. Gender, race and socioeconomic influence on diagnosis and treatment of thyroid disorders in the Brazilian Longitudinal Study of Adult Health (ELSA-Brasil). Braz J Med Biol Res. 2015;48(8):751–758. doi:10.1590/1414-431X20154445

15. “WHO Mean Body Mass Index (BMI)”. World Health Organization. Retrieved 5 February 2019

16. Wiersinga WM. De interpretatie van de bepaling van thyreoïdstimulerend hormoon (TSH) [The interpretation of the thyroid stimulating hormone (TSH) assay]. Ned Tijdschr Geneeskd. 2003 Jun 14;147(24):1156-8. Dutch. PMID: 12845833.

17. Fei X, Xing M, Wo M, Wang H, Yuan W, Huang Q. Thyroid stimulating hormone and free triiodothyronine are valuable predictors for diabetic nephropathy in patient with type 2 diabetes mellitus. Ann Transl Med. 2018;6(15):305. doi:10.21037/atm.2018.07.07

18. Asmelash D, Tesfa K, Biadgo B. Thyroid Dysfunction and Cytological Patterns among Patients Requested for Thyroid Function Test in an Endemic Goiter Area of Gondar, North West Ethiopia. Int J Endocrinol. 2019 Aug 14;2019:9106767. doi: 10.1155/2019/9106767. PMID: 31511773; PMCID: PMC6710807

19. Bharadiya AA, Aundhkar SC, Jaju JBet. Evaluation of thyroid function tests in patients with uncontrolled type 2 diabetes mellitus. Int J Health Sci Res. 2014;4(5):129–138.

20. Elmenshawi IM, Alotaibi SS, Alazmi AS, Prevalence of thyroid dysfunction in diabetic patients. J Diabetes Metab Disord Control. 2017;4(2):55–60. DOI: 10.15406/jdmdc.2017.04.00106

21. Ghazali and Abbiyesuku, Thyroid dysfunction in type 2 diabetics seen at the University College Hospital, Ibadan, Nigeria, Nig. J, Physiol. Sci. 25 (2010): 173–179 www.njps.physocnigeria.org

22. Brenta G. Diabetes and thyroid disorders. The British Journal of Diabetes & Vascular Disease. 2010;10(4):172–177. doi:10.1177/1474651410371321

23. Elgazar EH, Esheba NE, Shalaby SA, Mohamed WF. Thyroid dysfunction prevalence and relation to glycemic control in patients with type 2 diabetes mellitus. Diabetes Metab Syndr 2019;13:2513–7)

24. Tunbridge, D. C. Evered, and R. Hall, “The spectrum of thyroid disease in a community: the Whickham survey,” Clinical Endocrinology, vol. 7, no. 6, pp. 481–493, (1977)

25. Diez JJ, Sanchez P, Iglesias P. Prevalence of thyroid dysfunction in patients with type 2 diabetes. Exp Clin Endocrinol Diabetes 2011;119:201–7

26. Osei Sarfo-Kantanka, Fred Stephen Sarfo, Eunice Oparebea Ansah, Ishmael Kyei, “The Effect of Thyroid Dysfunction on the Cardiovascular Risk of Type 2 Diabetes Mellitus Patients in Ghana”, Journal of Diabetes Research, vol. 2018, Article ID 4783093, 8 pages, 2018. https://doi.org/10.1155/2018/4783093

27. Alsolami AA, Alshali KZ, Albeshri MA, Alhassan SH, Qazli AM, Almalki AS, et al. Association between type 2 diabetes mellitus and hypothyroidism: a case–control study. IntJGenMed. 2018;11:457–461 https://doi.org/10.2147/IJGM.S179205

28. Ogbonna SU and Ezeani IU Risk Factors of Thyroid Dysfunction in Patients With Type 2 DiabetesMellitus. Front. Endocrinol. 10:440. doi: 10.3389/fendo.2019.00440

29. Mehalingam V, Sahoo J, Bobby Z, Vinod KV. Thyroid dysfunction in patients with type 2 diabetes mellitus and its association with diabetic complications. J Family Med Prim Care 2020;9:4277–81.

30. Standard of care diabetes 2020 Diabetes Care 2020;43(Suppl. 1):S1–S2 | https://doi.org/10.2337/dc20-SINT

31. Martin SS, Daya N, Lutsey PL,. Thyroid Function, Cardiovascular Risk Factors, and Incident Atherosclerotic Cardiovascular Disease: The Atherosclerosis Risk in Communities (ARIC) Study. J Clin Endocrinol Metab. 2017;102(9):3306–3315. doi:10.1210/jc.2017-00986

32. Bernadette Biondi, George J Kahaly, R Paul Robertson, Thyroid Dysfunction and Diabetes Mellitus: Two Closely Associated Disorders, Endocrine Reviews, Volume 40, Issue 3, June 2019, Pages 789–824, https://doi.org/10.1210/er.2018-00163

